# Uncovering relationships between 24-hour rest-activity patterns and immune-metabolic dysfunction in young people with mood disorders

**DOI:** 10.1101/2025.08.26.24311025

**Authors:** Sarah McKenna, Mirim Shin, Shin Ho Park, Alissa Nichles, Natalia Zmicerevska, Connie Janiszewski, Minji Park, Elizabeth Phung, Frank Iorfino, Mathew Varidel, Elizabeth M Scott, Joanne S Carpenter, Jacob J Crouse, Ian B. Hickie

## Abstract

**Background:** Individuals with mood disorders are at increased risk of metabolic diseases such as Type 2 diabetes. Circadian (and linked 24 hour rest-activity) disturbances are highly prevalent among this population and have also been linked to immune-metabolic dysfunction. Currently there is limited understanding of the extent to which these pathophysiological processes co-occur across the various clinical stages of major mood disorders.

**Methods:** 225 young people (67% female; aged 23.65 ± 5.73 years) recruited from early intervention mental health services were assigned clinically to either Stage 1a or 1b (subthreshold disorders) or Stage 2+ (full threshold disorder) of illness according to the transdiagnostic staging model. We explored relationships between immune-metabolic risk factors (BMI, fasting glucose and insulin, insulin resistance, and CRP) and rest-activity parameters from actigraphy (24 hour ambulatory motor activity monitoring) using pairwise correlations, multiple linear regression interaction effects and subgroup analyses.

**Results:** For all participants, higher intradaily variability (greater rest-activity fragmentation) was associated with higher BMI (r=0.187, p=0.043), fasting insulin (r=0.180, p=0.031), HOMA2-IR (r=0.187, p=0.043), and CRP (r=0.178, p=0.032) across all stages of illness. Lower relative amplitude of rest-activity patterns indicating dampened circadian rhythmicity, was associated with higher BMI (β=-33.149, p=0.013) and CRP (β=-22.042, p=0.053), only for those in stage 2+ of illness. It was also associated with fasting insulin during stage 1b (β=-10.299, p=0.044) and stage 2 (β=-10.411, p=0.037), and with HOMA2-IR at stage 1b (β=-1.133, p=0.040). Finally, increased moderate-to-vigorous physical activity (MVPA) was associated with lower BMI only for those at Stage 2+ (β=-0.056, p=0.001).

**Conclusions:** Objective measures of blunted and fragmented 24 hour rest-activity (circadian) rhythms were associated with adverse immune-metabolic outcomes. Stabilisation and amplitude-boosting of rest-activity rhythms may be particularly valuable targets for indicated prevention and early intervention of major mood disorders.

## INTRODUCTION

Mood disorders, including bipolar, unipolar, and anxiety-depressive disorders are associated with 5 to 18 years reduced life expectancy, largely due to increased risk of premature cardiovascular disease (CVD). This risk is particularly pronounced in bipolar disorder (5-fold increase) and unipolar and anxiety-depressive disorders (3-fold increase).^1,2^ Thus, significant attention has been given to improved early detection and intervention of metabolic dysfunction, as it is a major risk to CVD risk.^3,4^ While weight gain and smoking are traditionally viewed as the main modifiable risk factors, lifestyle and dietary interventions show limited benefits for metabolic outcomes in this population, even achieving when they lead to weight loss.^5–7^ Moreover, genetic and large cohort longitudinal studies have implicated more complex pathophysiological processes, such as circadian disturbance, that may link mood disorders to increased cardiometabolic dysfunction.^8–10^ Further exploration of these processes may help to identify targets for monitoring, early intervention, and secondary prevention.

The circadian system is a biological mechanism that orchestrates physiological processes through intrinsic 24-hour light-dark cycles. This internal timekeeping system coordinates key metabolic processes, including glucose metabolism, insulin secretion, glucose tolerance, lipid regulation, energy expenditure, and appetite control.^11,12^ Disruption of circadian rhythms is associated with poor cardiometabolic outcomes, including metabolic syndrome and premature CVD. ^13,14^ This is evident in shift workers, whose irregular rest-activity cycles are associated with higher insulin resistance, plasma glucose and triglycerides.^15,16^ Furthermore, rest-activity cycle abnormalities such as delayed sleep phase, prolonged sleep period and excessive daytime inactivity, or high variability in daily timing or sleep onset and offset, are associated with mood disorders, particularly bipolar and atypical depressive subtypes^17–19^ , and have also been linked to more severe symptoms.^20–22^

Emerging evidence has shown that psychological (e.g., cognitive behaviour therapy for insomnia) and pharmacological (e.g., melatonin) therapies targeting sleep improve metabolic outcomes (e.g., weight stabilization and reduced insulin sensitivity) for individuals with emerging mood disorders,^21–25^ However, it is not yet clear which aspects of the rest-activity cycle are most important to address. For example, studies in pre-diabetic populations indicate that 24-hour circadian patterns, measured through actigraphy parameters like inter-daily stability, relative amplitude, and higher average activity level during the least active 5-h period (L5 ), show stronger association with metabolic outcomes (such as hemoglobin A1c [HbA1c], body mass index [BMI], and fasting glucose) than parameters related to sleep, such as sleep efficiency and sleep quality.^26–28^ Accordingly, circadian therapies that target rest-activity rhythms, for example by encouraging more activity in the morning as compared to the evening, will presumably be more effective at improving metabolic outcomes than traditional sleep therapies that focus on sleep quality or duration. ^26–28^ Improved understanding of the relationships between rest-activity rhythms and metabolic outcomes in individuals with mood disorders, will lead to more targeted assessment and intervention approaches in this population.

Two limitations in the current evidence base are the reliance on (a) markers that may be insensitive to early metabolic dysfunction, and (b) broad diagnostic categories that overlook illness stage (along which metabolic dysregulation and its predictors may vary). Young people aged 16 to 30 presenting to early intervention mental health services show heightened homeostatic model assessment of insulin resistance ([HOMA2-IR] (a measure of insulin sensitivity based on fasting insulin and glucose) despite normal rates of smoking and overweight.^29–31^ This metabolic dysfunction is also associated with elevated pro-inflammatory cytokines, including C-reactive protein (CRP), inerleukin-6 and tumour necrosis factor-alpha, which contribute to chronic inflammation and increased CVD risk.^32–34^ We previously compared immune-metabolic dysfunction in youth in in-patient hospital settings with more severe mental disorders^29^, and youth accessing ambulatory care, and showed those in inpatient settings with more severe disorders, had higher HOMA2-IR levels.^29^ In this population, age and BMI together accounted for only 22% of the variance in HOMA2-IR. ^29^ This warrants further exploration of physical health comorbidities at varying stages of illness.

The transdiagnostic clinical staging model offers a valuable framework for understanding these relationships, positioning individuals with mental illness along a continuum from, mild to moderate subthreshold symptoms (stages 1a and 1b) to more severe and complex disorders (stage 2+).^35–37^ These models can improve understanding of mental illness trajectories, including the emergence or progression of physical health comorbidities, and so can be used to understand how pathophysiological processes such as circadian disruption and metabolic dysfunction are linked to mood disorder severity.

Accordingly, the objective of the current study was to explore cross-sectional associations between objective (actigraphic) estimates of rest-activity and circadian motor activity rhythms and immune-metabolic markers (including HOMA2-IR and CRP) in young people with emerging mood disorders, seeking mental health care. We hypothesised that disturbed rest-activity patterns, in particular circadian rhythm disruptions, would be associated with metabolic risk factors, especially elevated HOMA2-IR. Given that both circadian disturbances and immune-metabolic dysfunction are associated with more severe mood disorders, we also hypothesized that these relationships would be heightened for those at a later stage of illness.

## METHODS AND MATERIALS

Participants were recruited between October 2004 and January 2024 to one of two transdiagnostic research studies at the University of Sydney’s Brain and Mind Centre investigating the neurobiology of emerging mental disorders in young people: the Youth Mental Health Follow Up Study (YMH) (Lee et al., 2018) and the Neurobiology Youth Follow Up Study (Nichles et al., 2021). The research was approved by the University of Sydney Human Research Ethics Committee (2012/1631) and the Human Research Ethics Committee of the Sydney Local Health District (2020/ETH01272), and all participants gave written informed consent, with parental/guardian consent obtained for participants younger than 16 years.

### Cohort selection

Study participants were drawn from a large cohort study of young people presenting to one of the youth mental health clinics associated with the University of Sydney’s Brain and Mind Centre, including early intervention services (*headspace*, Camperdown) and a private multidisciplinary clinic (Mind Plasticity), primarily serve young people with emerging mood or anxiety disorders.^38,39^ This cohort includes young people presenting to one of these services for mental health care who underwent blood assessments between November 2008 and July 2024. Individuals were included in the current study if they had undergone an actigraphy assessment within six months of undergoing a blood test to assess metabolic and immune blood markers. Participants were excluded from either study for insufficient English language skills to comprehend the protocol or clinically evident intellectual disability.

### Transdiagnostic clinical staging

Participants were assigned to clinical stages based on the transdiagnostic clinical staging model.^36,37,40^ This model places individuals on a continuum from early phases of illness with non-specific clinical presentations (Stage 1a ‘seeking help’) to those at greater risk with more specific subthreshold presentations (Stage 1b ‘attenuated syndromes’), or those who have already reached a threshold for a progressive or recurrent disorder (Stage 2, 3 or 4). Individuals in our sample were assigned to one of three groups (Stage 1a, Stage 1b or Stage 2+).^35^ We have reported substantial concordance between raters assigning young people to clinical stages (kappa = 0.72).^41^

### Sleep-wake and circadian variables

The 24-hour sleep-wake and circadian rest-activity parameters were measured using actigraphy recordings. Participants were instructed to wear an actigraph (GENEActiv Sleep device; Activinsights, Kimbolton, UK) on their non-dominant wrist for at least 14 consecutive days. The GENEActiv devices have been validated against several types of accelerometry-based activity monitors^42,43^ as well as for sleep–wake scoring^43^. Activity data was collected at 30 or 50hz (difference due to protocol updates).

Raw GENEActiv actigraphy data was processed using an open-source R package, GGIR (version 3.1.2) developed to process multi-day accelerometer data ^44^. Post processing was completed using the package mMARCH.AC (version 2.9.4) according to protocols established for the Mobile Motor Activity Research Consortium for Health (mMARCH) ^45^. Any days with less than 8 hours of data were removed. Missing data on remaining days was imputed with the mean of that epoch on all other days. Sleep Regularity Index (SRI) was calculated using the R package sleepreg (version 1.3.5) ^46^ based on sleep-wake estimates by epoch from GENEActiv GGIR output. A minimum of 5 overlapping days was required to calculate the SRI score.

Actigraphy parameters that were included in the current study were indicators of both sleep-wake and circadian rest-activity patterns.

Sleep-wake parameters included : Sleep midpoint (halfway point between sleep-onset and sleep-offset), total sleep time (number of minutes estimated as asleep per 24-hour period), sleep efficiency (percentage of time estimated as asleep between sleep onset and wake time), differences in weekend-weekday sleep midpoint, sleep regularity index (consistency of sleep and wake patterns over multiple days).

Rest-activity parameters include: Interdaily stability ([IS], a measure of how consistent an individual’s activity patterns are from day to day), intradaily variability ([IV], a measure of how often and how much a person’s activity levels change throughout the day), relative amplitude ([RA], the difference between the most active 10 hours (M10) and least active 5 hours (L5) by the formula (M10-L5)/(M10+L5)), moderate-to-vigorous physical activity ([MVPA], sum of one-minute epochs in which gross motor activity was larger than 100mg), and start time of the most active 10 hours.

### Metabolic, immune and other hormonal measures

Height and weight were collected by direct measurement or self-report for calculation of BMI using the formula: weight (kg) ÷ height (m^2^). Blood samples were collected in a fasting state at a pathology laboratory. The current analysis focused on blood markers for metabolic dysfunction (fasting plasma glucose, fasting insulin, cholesterol, triglycerides) and systemic inflammation (C-reactive protein). HOMA2-IR was calculated using fasting blood glucose and insulin with the HOMA2-IR (Homeostatic Model Assessment for Insulin Resistance) software V.2.2.3.^47^

### Statistical analysis

Statistical analyses were conducted in RStudio using R (version 4.2.3)^48^ between August, 2024 and November, 2024. Descriptive analyses were first used to provide an overview of demographic, actigraphy, clinical, and immune-metabolic data. Residual analyses were then conducted to verify that the conditions for linear regression had been met as regards direct relationships between immune-metabolic variables (BMI, fasting glucose, fasting insulin, HOMA2-IR, and CRP) and actigraphy variables.^49,50^ Subsequently, extreme outliers were deleted before conducting linear regression (threshold ±2 standard deviations; number of outliers deleted ranged from 4 to 12 (*M* = 6) across 15 analyses) to improve data distribution. Continuous variables were also centred to reduce skewness and to avoid multicollinearity when exploring interaction effects that could affect model convergence or inflate standard errors. ^49,50^ During subsequent analyses, missing data was handled using pairwise deletion so as to preserve more data as compared to listwise deletion.

Pearson correlation coefficients were used to assess bivariate relationships between actigraphy and immune-metabolic variables whilst controlling for age and gender. For all significant bivariate relationships, we explored interaction effects between clinical stage and actigraphy variables, to understand whether actigraphy variables account for a unique portion of the variance in immune-metabolic variables and whether these relationships vary between late as compared to early stages of illness. Linear regression was used to test interaction effects whilst controlling for age and gender.

Subsequently, interaction effects were also visually inspected using plots. We then used subgroup analyses to further explore relationships between actigraphy variables and immune-metabolic variables at each stage of illness.

## RESULTS

### Sample Characteristics

Participants were 225 young people (67.0% female; age 23.65 ± 5.73 years) attending early intervention mental health services. Clinical staging assessment categorized participants into three groups: Stage 1a (n=40; 47.6% female; age 22.98 ± 5.73), Stage 1b (n=143; 72.0% female; age 23.27 ± 5.59), and Stage 2+ (n=42; 67.5% female; age 25.53 ± 5.91, Table 1).

**Table 1.**
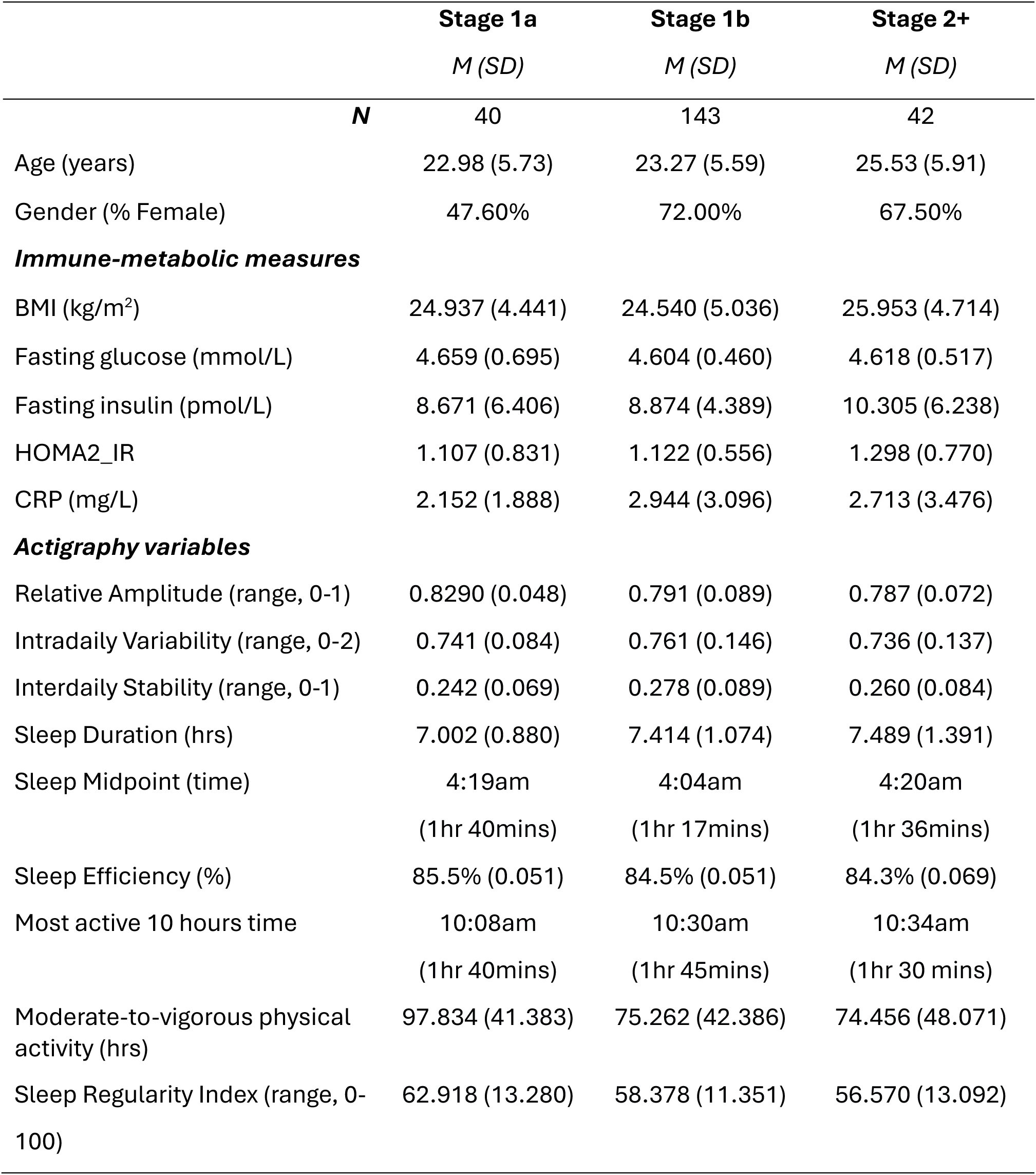
Descriptive statistics for demographic, immune-metabolic and actigraphy variables at each stage of illness.

|  | **Stage 1a** | **Stage 1b** | **Stage 2+** |
| --- | --- | --- | --- |
|  | *M (SD)* | *M (SD)* | *M (SD)* |
| **N** | 40 | 143 | 42 |
| Age (years) | 22.98 (5.73) | 23.27 (5.59) | 25.53 (5.91) |
| Gender (% Female) | 47.60% | 72.00% | 67.50% |
| ***Immune-metabolic measures*** |  |  |  |
| BMI (kg/m<sup>2</sup>) | 24.937 (4.441) | 24.540 (5.036) | 25.953 (4.714) |
| Fasting glucose (mmol/L) | 4.659 (0.695) | 4.604 (0.460) | 4.618 (0.517) |
| Fasting insulin (pmol/L) | 8.671 (6.406) | 8.874 (4.389) | 10.305 (6.238) |
| HOMA2_IR | 1.107 (0.831) | 1.122 (0.556) | 1.298 (0.770) |
| CRP (mg/L) | 2.152 (1.888) | 2.944 (3.096) | 2.713 (3.476) |
| ***Actigraphy variables*** |  |  |  |
| Relative Amplitude (range, 0-1) | 0.8290 (0.048) | 0.791 (0.089) | 0.787 (0.072) |
| Intradaily Variability (range, 0-2) | 0.741 (0.084) | 0.761 (0.146) | 0.736 (0.137) |
| Interdaily Stability (range, 0-1) | 0.242 (0.069) | 0.278 (0.089) | 0.260 (0.084) |
| Sleep Duration (hrs) | 7.002 (0.880) | 7.414 (1.074) | 7.489 (1.391) |
| Sleep Midpoint (time) | 4:19am<br>(1hr 40mins) | 4:04am<br>(1hr 17mins) | 4:20am<br>(1hr 36mins) |
| Sleep Efficiency (%) | 85.5% (0.051) | 84.5% (0.051) | 84.3% (0.069) |
| Most active 10 hours time | 10:08am<br>(1hr 40mins) | 10:30am<br>(1hr 45mins) | 10:34am<br>(1hr 30 mins) |
| Moderate-to-vigorous physical activity (hrs) | 97.834 (41.383) | 75.262 (42.386) | 74.456 (48.071) |
| Sleep Regularity Index (range, 0-100) | 62.918 (13.280) | 58.378 (11.351) | 56.570 (13.092) |

### Post-hoc power analysis to determine adequate sample size

Post-hoc power analysis indicated adequate power to detect medium to large effects (*r* = 0.015) when comparing groups (Stage 1a vs Stage 2+; power = 0.77); Stage 1b vs. other stages (power = 0.98). The analysis assumed a significance level of α = 0.05)

### Direct relationships between immune-metabolic and actigraphy variables

Correlation analyses revealed significant associations between circadian rhythm parameters and metabolic outcomes (Table 2). BMI was significantly positively associated with IV (r = 0.183, p = 0.043), indicating that more fragmented (rather than stable) rhythms within the 24hr day, were associated with higher BMI.

**Table 2.** Pairwise comparisons (Pearson correlation coefficients) between immune-metabolic and actigraphy variables controlling for age and gender.

|  | 1 | 2 | 3 | 4 | 5 | 6 | 7 | 8 | 9 | 10 | 11 | 12 | 13 |
| --- | --- | --- | --- | --- | --- | --- | --- | --- | --- | --- | --- | --- | --- |
| **1. BMI** |  |  |  |  |  |  |  |  |  |  |  |  |  |
| **2. Glucose** | 0.013 |  |  |  |  |  |  |  |  |  |  |  |  |
| **3. Insulin** | 0.433** | 0.366** |  |  |  |  |  |  |  |  |  |  |  |
| **4. HOMA2_IR** | 0.424** | 0.419** | 0.998** |  |  |  |  |  |  |  |  |  |  |
| **5. CRP** | 0.370** | 0.022 | 0.203* | 0.199* |  |  |  |  |  |  |  |  |  |
| **6. Relative amplitude** | -0.100 | -0.039 | -0.203* | -0.201* | -0.273** |  |  |  |  |  |  |  |  |
| **7. Intradaily variability** | 0.183* | 0.022 | 0.176* | 0.178* | 0.132 | -0.561** |  |  |  |  |  |  |  |
| **8. Interdaily stability** | 0.072 | 0.086 | 0.007 | 0.012 | 0.000 | 0.438** | -0.388** |  |  |  |  |  |  |
| **9. Sleep duration** | -0.009 | -0.022 | 0.033 | 0.033 | 0.149 | -0.079 | -0.028 | 0.013 |  |  |  |  |  |
| **10. Sleep midpoint** | 0.017 | -0.065 | -0.023 | -0.026 | -0.121 | -0.037 | 0.043 | -0.226** | -0.165* |  |  |  |  |
| **11. M10time<sup>1</sup>** | -0.052 | -0.029 | 0.023 | 0.022 | -0.004 | -0.389** | 0.189* | -0.415** | 0.046 | 0.698** |  |  |  |
| **12. MVPA<sup>2</sup>** | -0.074 | 0.058 | -0.097 | -0.088 | -0.224* | 0.711** | -0.466** | 0.294** | -0.175* | -0.120 | -0.296** |  |  |
| **13. Sleep efficiency** | -0.001 | 0.006 | -0.049 | -0.042 | 0.005 | 0.241** | -0.129 | 0.067 | 0.250** | -0.099 | -0.175* | 0.069 |  |
| **14. Sleep regularity index** | 0.100 | -0.009 | -0.081 | -0.076 | -0.146 | 0.383** | -0.040 | 0.193* | -0.067 | -0.147 | -0.226* | 0.140 | 0.515** |

**Note.** \*\*\* $p < 0.001$ ; \*\* $p < 0.01$ ; \* $p < 0.05$ . <sup>1</sup>MVPA = Moderate to vigorous physical activity; <sup>2</sup>M10time = Most active ten hours

Fasting blood glucose was not significantly correlated with any of actigraphy variables.

Fasting insulin was significantly negatively associated with RA (r = -0.203, p = 0.013) and positively associated with IV (r = 0.176, p = 0.031). This suggests that a more pronounced rest-activity rhythm across the 24hr day, as shown through higher RA, and more stable rhythms within days, shown through lower IV, are both associated with lower fasting insulin.

Likewise, HOMA2-IR was significantly negatively associated with RA (r = -0.201, p = 0.014) and positively associated with IV (r = 0.178, p = 0.032). Again, more robust rest-activity rhythms, leading to higher RA and lower IV, were both associated with lower HOMA2-IR.

An inflammatory marker, CRP, was also negatively associated with RA (r = -0.273, p = 0.005) and moderate-to-vigorous physical activity (r = -0.224, p = 0.023), suggesting the more robust rhythms and higher activity levels were associated with lower levels of inflammation.

As shown in Table 2, we also explored whether other actigraphy variables including IS, sleep midpoint, total sleep time, sleep efficiency, sleep regularity index, differences in weekend-weekday sleep midpoint, and timing of the most active 10 hours, were correlated with immune-metabolic variables but found no significant relationships. As such, we did not explore potential interaction effects with these variables.

### Exploring variations in the relationship between actigraphy and immune-metabolic variables according to stage of illness

#### BMI

Regarding BMI, we found a significant interaction effect (see Table 3; β= -39.653, p = 0.004) with RA, when comparing Stage 1b and 2+. Visual inspection of the graph (see Figure 1A) suggested the direction of the relationship was negative for those in Stage 2+, and positive for those in Stage 1b, but subgroup analyses showed that RA accounted for a unique portion of the variance in BMI only for those in Stage 2+ (see Table 4; β = - 33.149, p = 0.013). Thus, higher RA, indicating a more robust 24-hour rest-activity rhythm, was associated with lower BMI in those at Stage 2+ of mental illness.

**Figure 1.**
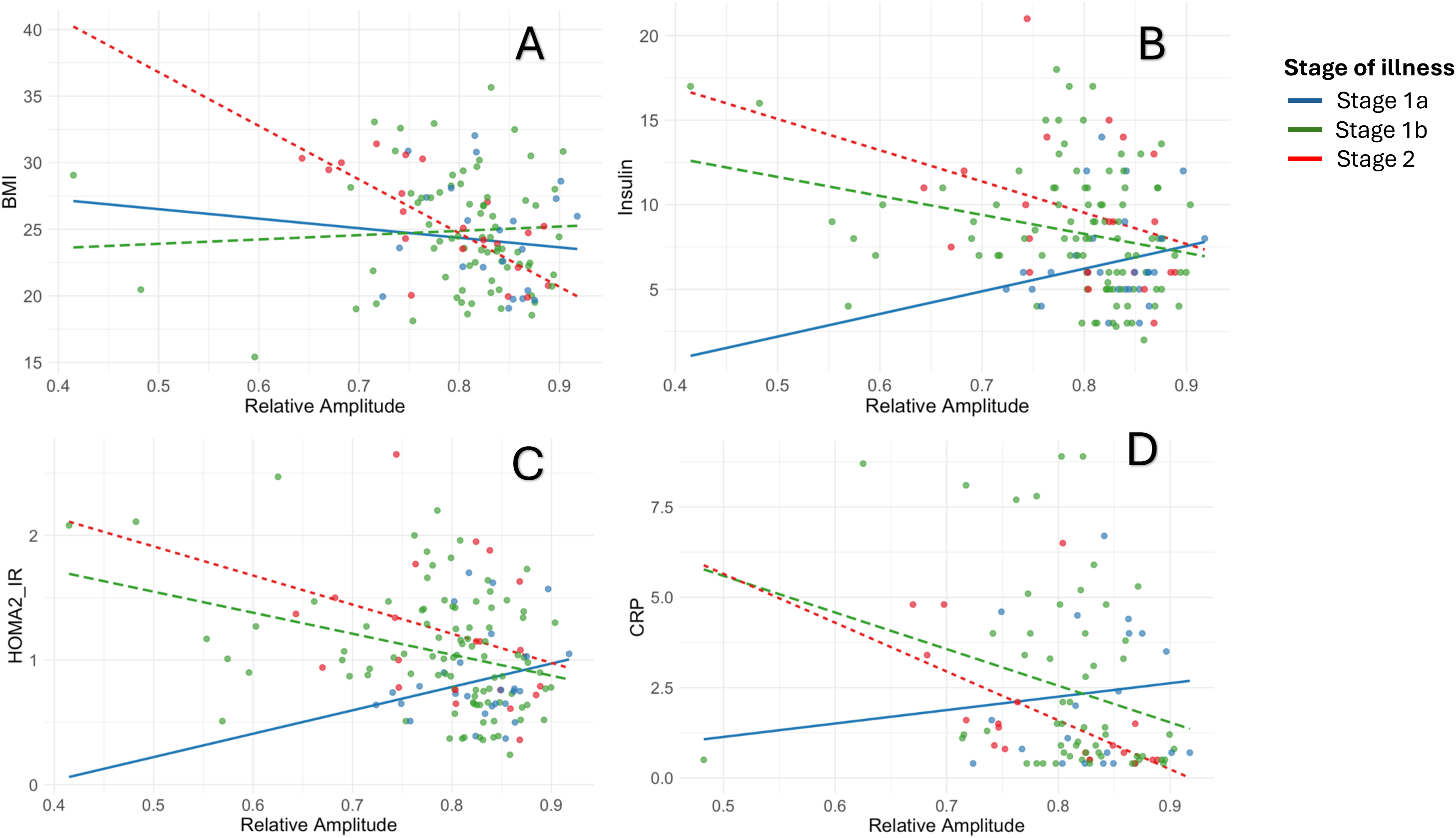
Comparing the relationship between relative amplitude and BMI (A), fasting insulin (B), HOMA2_IR (C), and CRP (D) between individuals at different clinical stages of mental disorders.

**Table 3.** Interaction effects (actigraphy * clinical stage) from multiple linear regressions exploring associations between actigraphy variables and immune-metabolic outcomes.

|  | Stage 1a vs. Stage 1b |  |  | Stage 1a vs. Stage 2 |  |  | Stage 1b vs. Stage 2 |  |  |
| --- | --- | --- | --- | --- | --- | --- | --- | --- | --- |
|  | *Estimate* | *Std. Error* | *p.value* | *Estimate* | *Std. Error* | *p.value* | *Estimate* | *Error* | *p.value* |
| **Relative amplitude** |  |  |  |  |  |  |  |  |  |
| *BMI* | 10.031 | 18.093 | 0.580 | -29.622 | 21.054 | 0.162 | **-39.653** | **13.392** | **0.004\*\*** |
| *Glucose* | -3.855 | 2.084 | 0.067 | -4.735 | 2.549 | 0.065 | -0.880 | 1.694 | 0.604 |
| *Insulin* | **-37.578** | **17.893** | **0.038\*** | **-48.235** | **21.733** | **0.028\*** | -10.658 | 14.897 | 0.476 |
| *HOMA2-IR* | **-5.100** | **2.272** | **0.026\*** | **-6.347** | **2.760** | **0.023\*** | -1.247 | 1.892 | 0.511 |
| *CRP* | -19.957 | 15.192 | 0.192 | **-37.174** | **16.773** | **0.029\*** | -17.217 | 9.804 | 0.083 |
| **Intradaily Variability** |  |  |  |  |  |  |  |  |  |
| *BMI* | 4.062 | 10.531 | 0.700 | 11.195 | 11.853 | 0.347 | 7.133 | 7.510 | 0.344 |
| *Glucose* | -1.599 | 1.199 | 0.185 | 1.744 | 1.392 | 0.213 | 0.145 | 0.868 | 0.868 |
| *Insulin* | -12.118 | 10.365 | 0.244 | -10.327 | 12.720 | 0.418 | 1.790 | 8.529 | 0.834 |
| *HOMA2-IR* | 1.381 | 1.318 | 0.297 | -1.204 | 1.617 | 0.458 | 0.176 | 1.084 | 0.871 |
| *CRP* | 4.045 | 9.086 | 0.657 | -1.483 | 10.043 | 0.883 | -5.529 | 5.879 | 0.350 |
| **Moderate to vigorous physical activity** |  |  |  |  |  |  |  |  |  |
| *BMI* | 0.034 | 0.023 | 0.139 | -0.022 | 0.026 | 0.403 | **-0.056** | **0.022** | **0.010\*** |
| *Glucose* | -0.005 | 0.003 | 0.091 | 0.005 | 0.003 | 0.064 | -0.002 | 0.003 | 0.442 |
| *Insulin* | -0.007 | 0.023 | 0.752 | -0.040 | 0.028 | 0.161 | -0.033 | 0.023 | 0.162 |
| *HOMA2-IR* | -0.001 | 0.003 | 0.632 | 0.001 | 0.003 | 0.632 | -0.004 | 0.003 | 0.170 |
| *CRP* | -0.031 | 0.018 | 0.086 | -0.038 | 0.021 | 0.069 | -0.007 | 0.017 | 0.679 |

Note: BMI = body mass index; HOMA2-IR (Homeostatic Model Assessment of Insulin Resistance); CRP = C-reactive protein; MVPA = Moderate-to-vigorous physical activity

**Table 4.**
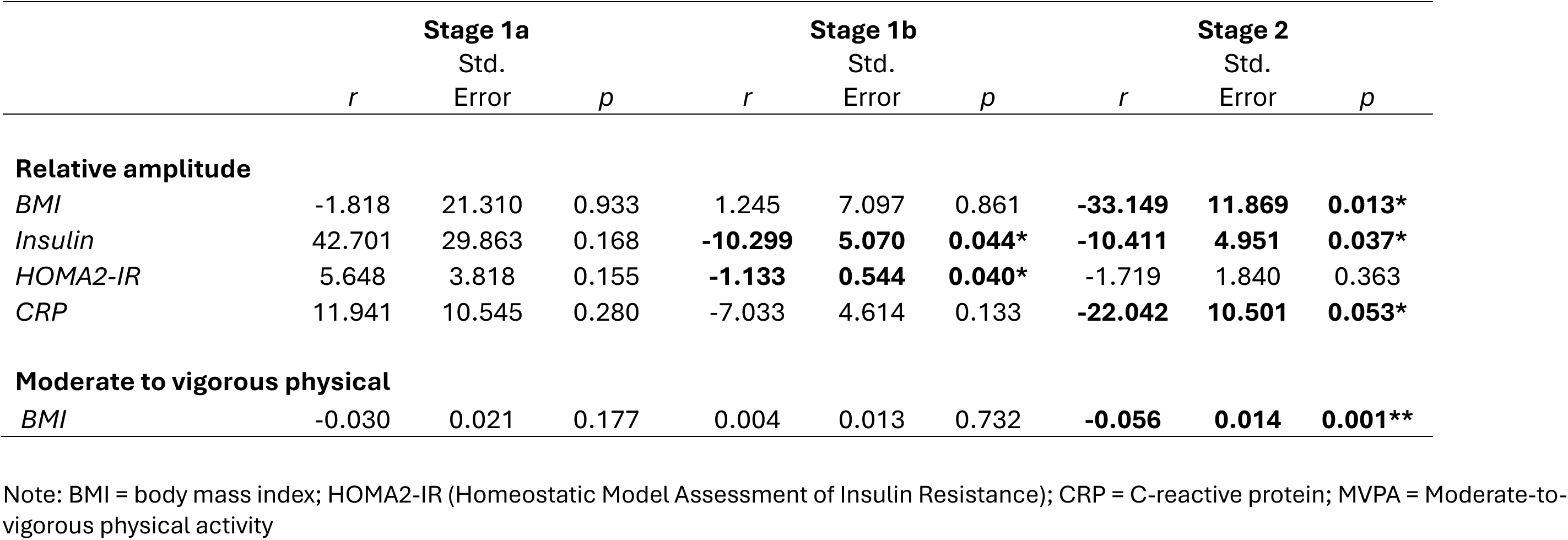
Subgroup analyses comparing direct relationships between actigraphy variables and immune-metabolic outcomes between stage of illness.

We also found a significant interaction effect for MVPA and clinical stage when comparing Stage 1b and Stage 2+ (see Table 3; β = -0.056, p = 0.010). Visual inspection of the graph suggested that the direction of the relationship between BMI and physical activity was positive for those in Stage 1b and negative for those in Stage 2+ (see Figure 2), but subgroup analysis showed that physical activity was uniquely associated with BMI only during Stage 2+ of illness (see Table 4; β = -0.056, p = 0.001). We did not find significant interaction effects when comparing relationship between BMI and IV (see Table 3).

**Figure 2.**
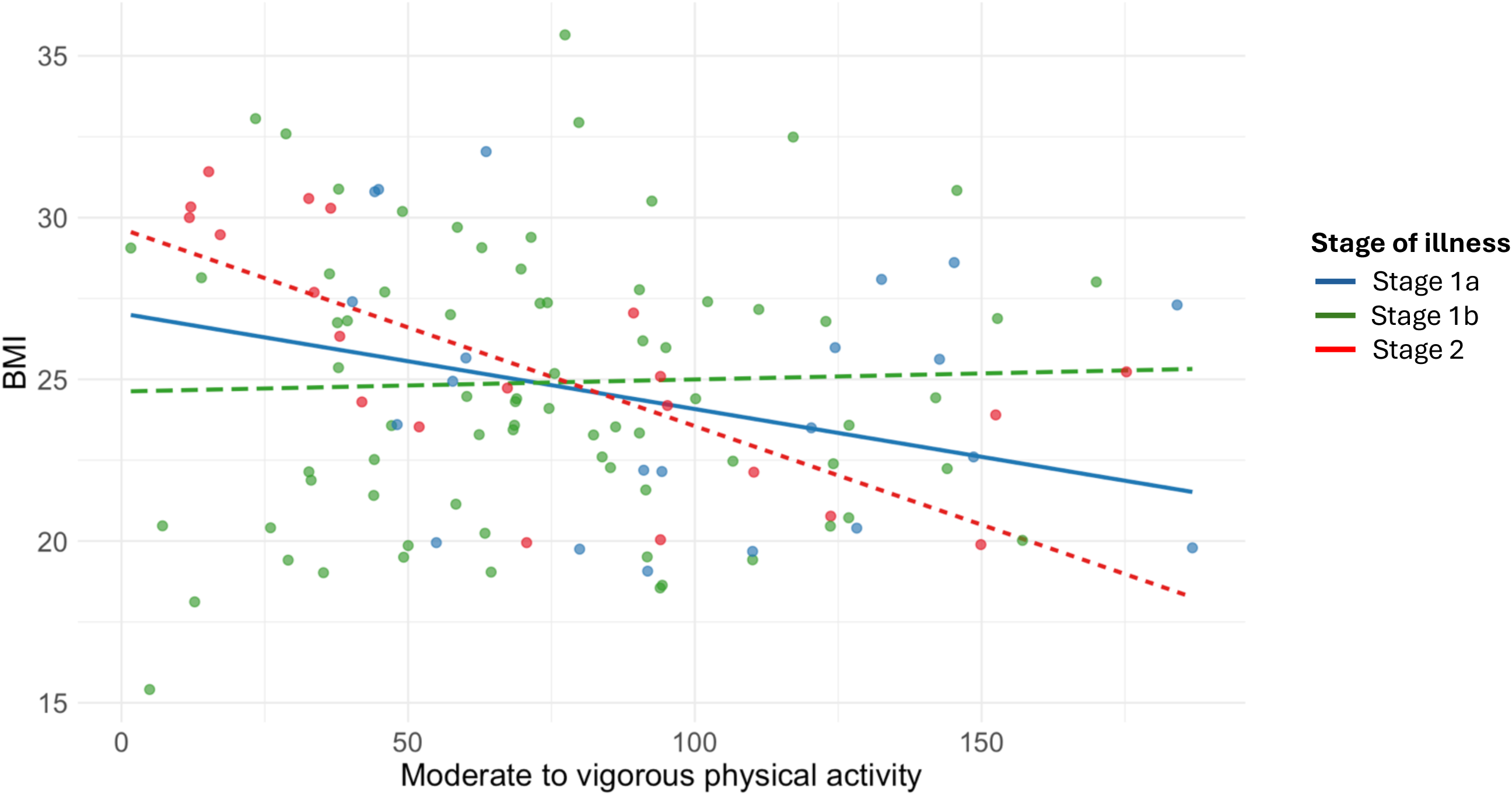
Comparing the relationship between moderate-to-vigorous physical activity (MVPA) and BMI between individuals at different clinical stages of mental disorders.

#### Fasting glucose

We did not find any significant interaction effects when looking at the relationship between fasting glucose and actigraphy variables (see Table 3).

#### Fasting insulin

When examining RA, we found two significant interaction effects, when comparing those in in Stage 1b ( β = -37.578, p = 0.038) and Stage 2+ ( β = -48.235, p = 0.028) to those in Stage 1a. Figure 1B suggested that the direction of the relationship between fasting insulin and RA was positive for those in Stage 1a and negative for those in Stage 1b and 2+; but subgroup analyses showed that RA accounted for a unique portion of variance in fasting insulin for those in Stage 1b ( β = -10.299, p = 0.044) and Stage 2+ ( β = -10.411, p = 0.037), not in Stage 1a. For both of these groups, higher RA was associated with lower fasting insulin. We did not find significant interaction effects when looking at IV or MVPA.

#### HOMA2-IR

When examining HOMA2-IR, we found an interaction effect when comparing Stage 1a with Stage 1b ( β = -5.100, p = 0.026) and Stage 2+ ( β = -6.347, p = 0.023). Similar to fasting insulin, visual inspection (Figure 1C) also showed positive relationship between HOMA2-IR and RA in stage 1a but negative relationship in Stage 1b and 2+.

However, RA accounted for a unique portion of variance in HOMA2-IR only for individuals in Stage 1b of mental illness ( β = -1.133, p = 0.040). Again, for this group, more robust 24-hour rest-activity rhythms (shown by higher RA) was linked to lower HOMA2-IR. We did not find significant interaction effects when looking at IV or MVPA.

#### CRP

For RA a significant interaction effect was found when comparing individuals in Stage 1a and Stage 2+ ( β = -37.174, p = 0.029). Visual inspection of the graphs (Figure 1D) suggested that there was a positive relationship between CRP and RA for those in Stage 1a of illness, and a negative relationship for those in Stage 2+. Subgroup analysis showed that RA accounted for a unique portion of the variance in CRP for those in Stage 2+ of illness only ( β = -22.042, p = 0.053). In this group, more pronounced rest-activity rhythms were associated with lower CRP. We did not find any significant interaction effects when exploring interaction effects associated with IV or MVPA (see Table 3).

## DISCUSSION

Better targeted indicate prevention and early intervention strategies s are needed to improve immune-metabolic outcomes for people with severe mood disorders. This requires a better understanding of the mechanisms driving poor physical health outcomes in this population. Here, we found cross-sectional evidence that the robustness and fragmentation of circadian rest-activity rhythms may be implicated in dysregulated metabolic outcomes. Many of these relationships were more pronounced in later stages of illness. Greater fragmentation of circadian rest-activity rhythms (IV) was related to higher BMI, fasting insulin, and HOMA2-IR at all stages of illness, whereas the robustness of rest-activity rhythms (RA) was uniquely associated with BMI, insulin, HOMA2-IR, and CRP only in later stages of illness, but not for those in Stage 1a. Moderate-to-vigorous physical activity was only related to BMI for those who had met the threshold for a severe mental disorder (Stage 2+). Sleep disturbances, including delayed sleep midpoint (ranging from 4:04am to 4:20am) as well as shorter sleep duration (7 hours to 7.5 hours), were found across all groups, consistent with previous studies in youth with mood disorders.^51,52^ Thus, sleep-wake disturbances are prevalent across all stages of mood disorders, but relationships to metabolic health may not appear until later in illness. Based on these cross-sectional findings, there is an urgent need to establish in prospective studies whether circadian disturbances are causally related to metabolic dysfunction for those with mood disorders.

A key finding is that rest-activity measures related to circadian patterns were significantly associated with BMI and insulin resistance, whereas estimates of sleep, including sleep duration, were not. By contrast, existing research from psychiatric populations has produced mixed findings. In patients with schizophrenia more fragmented and variable rest-activity rhythms were associated with higher HbA1c levels and worse physical health.^53^ Whereas, physical activity and sleep efficiency but not IS partially mediates the association between lifetime Atypical Major Depressive Disorder and BMI and metabolic syndrome.^54^ Likewise, in populations with Bipolar disorder in remission, fragmentation index but not RA was correlated with systolic blood pressure and atherogenic index of plasma, while a sleep-wake parameter, sleep efficiency, was correlated with metabolic outcomes including triglycerides.^55^ Importantly, much of this research is in adult populations with established disorders and has not assessed insulin sensitivity. We found that rest-activity variables were differentially related to the metabolic outcomes included in our research, and that relationships varied at different stages of illness, with physical activity more strongly related to BMI at later stages.

Given that biological circadian rhythms, such as core body temperature amplitude, have strong links to metabolite rhythms, there is impetus to conduct more detailed longitudinal research, particularly in youth cohorts, to gain a more nuanced understanding of relationships between circadian rhythm disruption and metabolic dysfunction in individuals with mood disorders.^18,56^ As well as supporting more targeted interventions for metabolic dysfunction, this approach could also lead to a better understanding of the pathophysiological processes underlaying trajectories to major mood disorders.

Another important finding is that while physical activity was associated with BMI this was only in later stages of illness (Stage 2+), whereas RA and IV were associated with fasting insulin and HOMA2-IR during earlier stages of illness. A potential explanation is that insulin resistance (including elevated fasting insulin and HOMA2-IR) may emerge earlier and be more strongly impacted by circadian disturbances than weight gain. This aligns with previous research from early intervention services showing that insulin resistance is a more sensitive early marker of emerging metabolic dysfunction than BMI or fasting glucose.^29–31^ By contrast, elevated BMI emerges later in illness, when individuals have been exposed to a complex mix of risk factors, such as medication and poor lifestyle, for an extended period of time. Taken together, there is increasing evidence that insulin resistance should be monitored in early intervention services in those at risk of later CVD, despite not being recommended by current guidelines.^57–59^

Despite these novel insights, the research has important limitations. Firstly, group sizes were not uniform across stage of illness, with Stage 1a and Stage 2 being far smaller (around 40 in each group) than Stage 1b. As such, we may have failed to detect significant relationships in these two groups. For this reason, further longitudinal research in larger cohorts is needed to explore patterns in metabolic and circadian variables across key stages of mental illness progression. Relatedly, given this study utilised cross-sectional data, we were unable to explore the direction of the relationship between actigraphy and metabolic outcomes. It is currently unclear whether the metabolic dysfunction and circadian disruptions observed here co-occur and are driven by the same underlying mechanisms, or whether they are causally related. Another important limitation is that metabolic variables assessed in the paper are themselves subject to circadian variation which we did not capture, more consistent measurements taken at various time points may give a better picture of links to circadian disturbance. Another measurement issue was that our study protocols involved collecting BMI both through direct measurement and self-report, we did not record which method was used. This may have introduced bias to our data analyses and could have influenced our exploration of BMI and actigraphy variables. Further, due to lack of data, we were unable to account for the potential effects of psychotropic medications. SSRI’’s and antipsychotic medications commonly used to treat mood disorders, have been linked to adverse metabolic outcomes including weight gain and increased risk of diabetes.58,59 Likewise, melatonergic medications, Benzodiazepines, and anti-psychotic medications have been shown to adversely affect circadian functioning.60,61 Our current protocols involve collecting more information about current medication, along with detailed metabolic and actigraphy data, this will allow us to properly account for the effects of medication in future analyses. Given the critical need to reduce the mortality gap for those with mood disorders, this creates an urgent imperative for more targeted research on rest-activity rhythms and their relationship to metabolic processes.

The current research was the first to investigate relationships between objective 24-hour rest-activity patterns and metabolic variables in youth attending early intervention services for mental healthcare. Parameters related to circadian rhythms, including relative amplitude, inter-daily stability, and moderate to vigorous physical activity were related to a range of metabolic and inflammatory outcomes including BMI, fasting insulin, HOMA2-IR, and CRP, whereas other sleep variables such as duration, midpoint, and efficiency were not associated with such outcomes. Moreover, relative amplitude and physical activity appeared to be more strongly related to metabolic outcomes at later stages of illness. Overall, boosting the amplitude and stability of circadian rhythms may be an important target for future interventions in youth with mood disorders^22^ and have important links to both mental and physical health outcomes.

## Data Availability

All data produced in the present study are available upon reasonable request to the authors

## Acknowledgements

The authors acknowledge the Gadigal people of the Eora nation, upon whose ancestral lands our research was conducted. We pay our respect to elders past and present. We thank all the young people who have participated in this study, and all the staff in the Youth Mental Health Team at the Brain and Mind Centre, past and present, who have contributed to this work.

